# Urban-Rural Disparities in Spatio-Temporal Accessibility of Pharmacy Care: A Case Study of Vermont, USA

**DOI:** 10.1101/2025.04.29.25326678

**Authors:** Samuel Roubin, Joseph Holler, Peter Kedron

**Affiliations:** Department of Geography, Middlebury College, Middlebury, VT 05753; Department of Geography, University of California Santa Barbara, Santa Barbara, California, 93106

**Keywords:** Pharmacy, spatial accessibility, Vermont, E2FSCA, rural healthcare, health equity, open-science, GIS

## Abstract

**Background:** Pharmacies are more accessible than other health care services, providing front-line primary and preventative health care. Recent research has underscored the importance of pharmacy access for reaching underserved and rural populations. However, minimal attention has been given to the spatial and temporal dimensions of access to pharmacy care despite the evidence suggesting that geographic accessibility impacts the utilization of various healthcare services.

**Methods:** In this study, we measured spatiotemporal variation in access to pharmacy care across Vermont using the Enhanced 2-step Floating Catchment Area (E2SFCA) method. We surveyed pharmacies during Fall 2023 in Vermont to collect data on staffing levels and hours of operation and paired this data with population and an OpenStreetMap road network. We computed hourly spatial accessibility scores by town geographic units across for weekdays, Saturdays, and Sundays, and compared results for metropolitan, micropolitan, and rural areas, testing for significance using the Kruskal-Wallis H Test.

**Results:** Our findings reveal significant disparities in pharmacy access between rural and urban areas across virtually all temporal segments. Accessibility was diminished during weekends across all locations, highlighting challenges in accessing care outside of conventional business hours. Accessibility gaps between rural and non-rural towns were exacerbated outside of standard weekday business hours.

**Conclusions:** Our study revealed important urban-rural differences in the spatial accessibility of pharmacies across Vermont and characterized how these accessibility differences vary temporally, with increasing disparities outside normal working hours. The results are useful for informing public health policy and the provision of health services. We have also demonstrated the importance of data on pharmacy operating hours and service levels for modeling spatiotemporal pharmacy accessibility, highlighting the need for further research and improving centralized databases to facilitate research in this field.

## Background

Pharmacy care has emerged as an increasingly important component of the healthcare landscape. Pharmacies perform a front-line role in public health by providing services such as medicine dispensing, patient consultation, preventative screenings, disease management, vaccine delivery, and diagnostic testing. These services reflect a continued shift from “product-centered” to “patient-centered” pharmacy care, where holistic and consultive services are becoming increasingly emphasized [1,2]. Recently, pharmacies have been instrumental in supporting the United States’ response and recovery throughout the COVID-19 pandemic [3]. Pharmacies are well-suited to provide preventive healthcare services due to their convenient locations, increased operational hours, and improved flexibility [4,5]. They also play a crucial role in ensuring equitable access to healthcare by reaching underserved individuals who lack access to other healthcare settings [5], as well as rural residents who are less frequently served by primary care providers [6]. However, recent restructuring in the pharmacy industry has created a significant trend of retail pharmacy closures across the United States, creating gaps in spatial accessibility with adverse impacts on rural and marginalized communities [7–11].

Despite the important and expanded front-line role that pharmacies play in public health, their spatial and temporal accessibility remains understudied [12]. Most existing studies treat pharmacy locations as homogenous, residential areas as centroids, and calculate distances as straight lines [12]. Spatial or geographic accessibility of healthcare services is increasingly recognized alongside other accessibility factors, such as affordability, availability, and acceptability, and measured using geographic information systems (GIS). Research indicates that geographic access impacts the utilization of healthcare services, including primary care [13] and hospitals [14], and that longer travel times to pharmacies are associated with lower utilization rates [15]. Despite this evidence, the literature on geographic access to pharmacies is limited and existing studies rely on density or distance measures that do not account for distance decay or spatial interaction between locations of supply and demand [12].

We address this gap in the literature by applying the Enhanced Two-Step Floating Catchment Area (E2SFCA) method [16] at one-hour intervals for typical weekdays and weekends for Vermont pharmacies. Two-step floating catchment (2SFCA) approaches to measuring spatial accessibility are widely used and improve upon simpler methods by modeling spatial interactions, measuring accessibility for a residential area of demand as the sum of service-to-population ratios for catchment areas around service locations [17,18]. The “enhanced” form (E2SFCA) also accounts for distance decay, modeling the decreasing likelihood of interaction between more distant locations [16].

To our knowledge, only four other studies have applied the 2SFCA [19–21] or E2SFCA [22] method to pharmacy access, and none of these considered how accessibility varies by time. Our study contributes to this literature by measuring varying levels of service at retail pharmacy locations and by adding a temporal dimension to accessibility. The limited use of floating catchment methods for studies of pharmacy access may be rooted in the lack of data on service levels and locations. All four floating catchment studies cited above treated pharmacies as equivalent in terms of levels of service or supply, whereas the floating catchment method requires a measure of service at each location. We address these limitations by surveying each pharmacy location in Vermont for their staffing levels and typical hours of operation. This unique combination of data allows us to ask three novel questions about spatio-temporal patterns of pharmacy access at underserved places and times in Vermont.

First, does spatial accessibility to pharmacies vary between metropolitan, micropolitan, and rural towns in Vermont during conventional business hours? Rural populations across the United States generally have more limited geographic access to pharmacies, with a smaller proportion of rural census tracts having access to at least one pharmacy per 10,000 people compared with suburban and urban tracts [21,23]. Additionally, rural populations disproportionately rely on a single “keystone” pharmacy for access and may be more severely impacted by pharmacy closures [10,24], which is particularly concerning given the recent widespread closures [8,9,11]. Importantly, Vermont’s population geography is predominantly rural, with just one metropolitan region and parts of four micropolitan regions within the state.

Second, does the accessibility of pharmacies vary temporally over different days of the week and times of day on typical weekdays, Saturdays, and Sundays? In addition to spatial inaccessibility, rural residents are less likely than their metropolitan counterparts to have access to primary care providers during nights and weekends [25], potentially exacerbating existing geographic limitations. Recognizing the impact of time on healthcare accessibility, some spatial accessibility analyses of healthcare have evolved to investigate the temporal dimension of access [26–28]. However, we are not aware of any studies that have examined the spatial accessibility of pharmacies in a temporally explicit manner, leaving a critical gap given the time-sensitive nature of many pharmacy services.

Third, if differences in pharmacy accessibility between rural and non-rural towns exist, are urban-rural differences in access exacerbated outside of conventional business hours? Urban-rural spatial accessibility gaps may be exacerbated outside of normal business hours, supported by results from spatio-temporal access to pediatrics in Taiwan [29] and general practitioner surgeries in Whales [30]. Furthermore, rural patients seek after-hours urgent care less frequently than urban patients due to distance barriers [31,32] and bypass nearby clinics to seek primary care at more distant clinics with after-hours options in New Zealand [33].

## Methods

This descriptive study has been conducted following open science practices with a registered pre-analysis plan, a compendium of protocols, data and code, and a post-analysis report [34]. We analyzed data using the CyberGISX JupyterHub system [35] hosted at the University of Illinois Urbana-Champaign. This study extends the work of Kang and others’ study of COVID-19 care in Illinois [36] and includes improvements from Holler et al.’s reproduction and reanalysis [37] of that study. The following sections summarize the study’s geographic context, data, and methods, each of which differ substantially from Kang et al. [36].

### Geographic Context

Our study is focused on the state of Vermont. Vermont has the highest proportion of its residents living in rural areas of any state in the US, at 64.9% [38]. In addition to its predominantly rural population, Vermont also ranks as the state with the fourth highest proportion of people aged 65 and older [39], and its rural areas tend to be older than its urban areas. Older populations are more reliant on pharmacy services given their higher utilization rates of prescription medications compared to younger demographics [40]. Simultaneously, elderly populations experience limitations in mobility, making geographic accessibility especially important. A county-level survey of pharmacists in 2021 found the number of pharmacists was highest in urban areas (112.7 full-time equivalents per 100,000 people in Chittenden County) and lowest in rural areas, with two counties having no pharmacists [41].

Understanding gaps in spatial accessibility to pharmacies in Vermont is particularly crucial given these unique demographics.

We chose census county subdivisions (n = 255) as the spatial unit of analysis for the study because they are compatible with the administrative boundaries of incorporated towns and municipalities and the US Census New England City and Town Areas (NECTA) classifications. Vermont has one metropolitan area, Burlington, with 35 county subdivisions. Another 48 county subdivisions belong to four micropolitan areas centered on Rutland, Barre, and Bennington in Vermont, and Lebanon in New Hampshire. The remaining 172 of 255 county subdivisions in Vermont are rural. Clusters of pharmacies tend to appear in metropolitan and micropolitan areas with greater population densities, whereas pharmacies are sparsely distributed in rural areas across Vermont (Figure 1).

**Figure 1.**
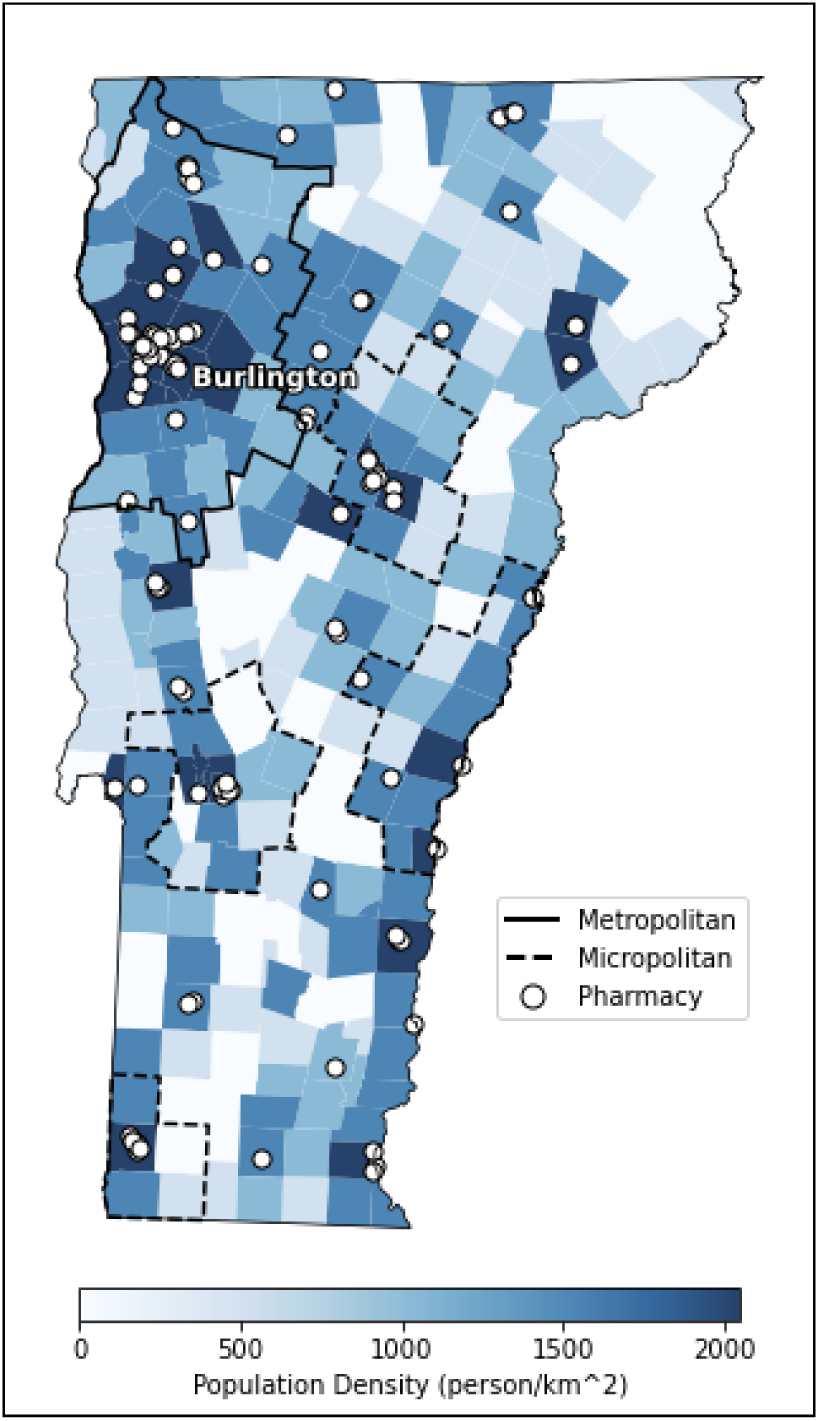
Vermont Study Area with population density across all towns (n = 255) and in-state pharmacy locations (n = 117). Metropolitan and micropolitan towns are outlined.

### Data Sources and Collection

Our temporally explicit E2SFCA model required three data inputs: 1) Retail pharmacy locations, including hours of operations and staffing levels, 2) county subdivisions with population totals, and 3) road networks.

Given the absence of centralized data on pharmacy staffing and hours of operation, we gathered this information using a three-step data collection process. First, we compiled a comprehensive spatial database of pharmacies inside and within 10 miles of Vermont with data from the Vermont Office of Professional Regulation (OPR) and the Department of Homeland Security Homeland Infrastructure Foundation-Level Data (HIFLD). We verified this data with OpenStreetMap (OSM) and Google Maps to ensure the completeness and currency of pharmacy location data, resulting in 117 locations inside Vermont and 75 locations in New Hampshire, Massachusetts, and New York. Second, we attempted to contact the store or regional managers for all 117 Vermont retail pharmacies under Middlebury College Institutional Review Board Protocol 264 between October and December of 2023, yielding responses for 102 locations (87%). Third, for the remaining 15 Vermont locations and 75 out-of-state locations, we utilized publicly available web pages to record hours of operation and imputed average staffing levels based on pharmacy type (independent or retail chain).

Population data at the county subdivision level was sourced from the United States Census Bureau American Community Survey (ACS) 2018-2022 (5-year estimates), along with metro/micropolitan classification from the Census NECTAs. We selected county subdivisions for this analysis because they offer a level of geographic detail comparable to census tracts in Vermont and align with the NECTA classifications and political units of town planning. Data for New York, New Hampshire, and Massachusetts was included to avoid boundary effects and ensure that all populations who use the pharmacies in our dataset were accounted for. We acquired and compiled data in R using TidyCensus [42].

We extracted and constructed a road network from OSM in Python using OSMnx [43], including roads up to 30 miles beyond Vermont. OSMnx functions were used to correct topological errors and impute any missing speed limit data.

### Enhanced Two-Step Floating Catchment Area (E2SFCA) Method

We utilized the Enhanced 2-Step Floating Catchment Area (E2SFCA) method [16] to descriptively assess spatial and temporal dimensions of pharmacy accessibility across Vermont. Our implementation of the E2SFCA measures the number of pharmacists available per 10,000 residents. The conventional two-step floating catchment area (2SFCA) method works in two main steps. In the first step, a service to population ratio *R*_*j*_ is calculated for each service location

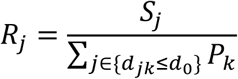

where *S*_*j*_ is the service capacity at location *j* and *P*_*k*_ is the population of the residential location *k*. The distance between each residential location and service location is *d*_*jk*_, and *d*_0_ is a constant distance threshold. In the second step, the accessibility *A*_*i*_ at each residential location *i* is calculated through the summation of all service-to-population ratios *R*_*j*_ (calculated in step 1) of all provider locations *j* that have a distance *d*_*ij*_ from the residential location within the threshold

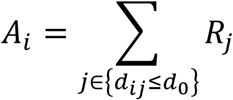

The enhanced method (E2SFCA) accounts for the decreasing likelihood of accessing services as distance increases, known as distance decay. Each catchment is broken into a set of segments *D* defined by constant distance ranges, and each range is assigned a corresponding weight *W*. The distances and weights are a discrete approximation of continuous distance decay, assigning lower weights to more distant places [16,18]. These weights are applied to both the population at each residential location and service to population ratio at each service location:

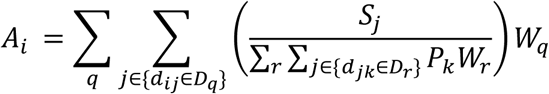

In this study, *S*_*j*_ is the level of services at each retail pharmacy location, estimated as the number of pharmacists on staff at a time plus one-half the number of pharmacist technicians. We added a temporal dimension to the accessibility analysis by calculating the E2SFCA model for each hour of a typical weekday, Saturday, and Sunday, thereby accounting for operating hours of pharmacy services and assuming consistent staffing levels at each location throughout the hours of a given day.

We defined *P*_*k*_ as the total population of county subdivisions. We use the same county subdivision geographic units for our final accessibility measure *A*_*i*_. Following precedents of prior studies [16,36], we defined three catchment distance segments *D* as a set of travel times (minutes): [0 to 10, 10 to 20, 20 to 30] with associated weights *W*: [1.0, 0.68, 0.22]. We calculated travel-times driving on roads at the speed limit using the NetworkX Python package and converted the network results into geographic polygon areas using the convex hull method.

Floating catchment studies typically simplify population areas (*P*_*k*_) into centroid point locations for the purpose of determine their distance relationship with services (*d*_*jk*_). This method increased uncertainty wherever a population area is only partially within a particular distance catchment *D*_*r*_. We implemented a more robust solution by overlaying the catchment (*D*_*r*_) with county subdivision polygons (*P*_*i*_) to measure the proportion of overlapping area and calculate an area-weighted population estimate. The population of a county subdivision assumed to be in a given catchment segment is proportional to the proportion of the county subdivision’s area covered by the catchment segment.

In sum, our implementation of a temporally explicit E2SFCA model yields accessibility scores for each Vermont county subdivision and hour of the day, interpretable as the number of retail pharmacists per 10,000 people likely to use pharmacies, given distance constraints.

### Statistical Analysis

We answered our first question on urban-rural disparities by focusing on a weekday time when all pharmacies are open, averaging town-level accessibility scores according to the metro-micro-rural classification and testing for significance with the Kruskal-Wallis *H* test (a non-parametric one-way analysis of variance test based on ranks). We answered our second question on temporal variability by first summarizing the maximum accessibility by weekday, Saturday, and Sunday for each county subdivision. We calculated the mean accessibility by day, mapped the spatial distribution each day, and tested for significance with the Kruskal-Wallis *H* test.

Finally, we answered our third question on whether urban-rural divides are exacerbated at off-hours by comparing the percent decline in service from weekdays to Saturdays and Sundays, as well as across hours of these days, for urban towns (metro and micro combined) and rural towns.

## Results

### Metropolitan, Micropolitan, and Rural Spatial Accessibility

We found that spatial accessibility to pharmacies varies considerably between metropolitan, micropolitan, and rural towns in Vermont during conventional business hours (Figure 2), with higher mean peak accessibility in metropolitan and micropolitan areas than rural areas. The mean accessibility scores were 4.20, 4.25, and 3.01 for metropolitan, micropolitan, and rural areas, respectively, with significant differences between them (Kruskal-Wallis H = 14.2, *p* < 0.001). Micropolitan areas had virtually the same access as metropolitan areas (1% difference), while rural areas had 28% less access on average compared to metropolitan areas.

**Figure 2.**
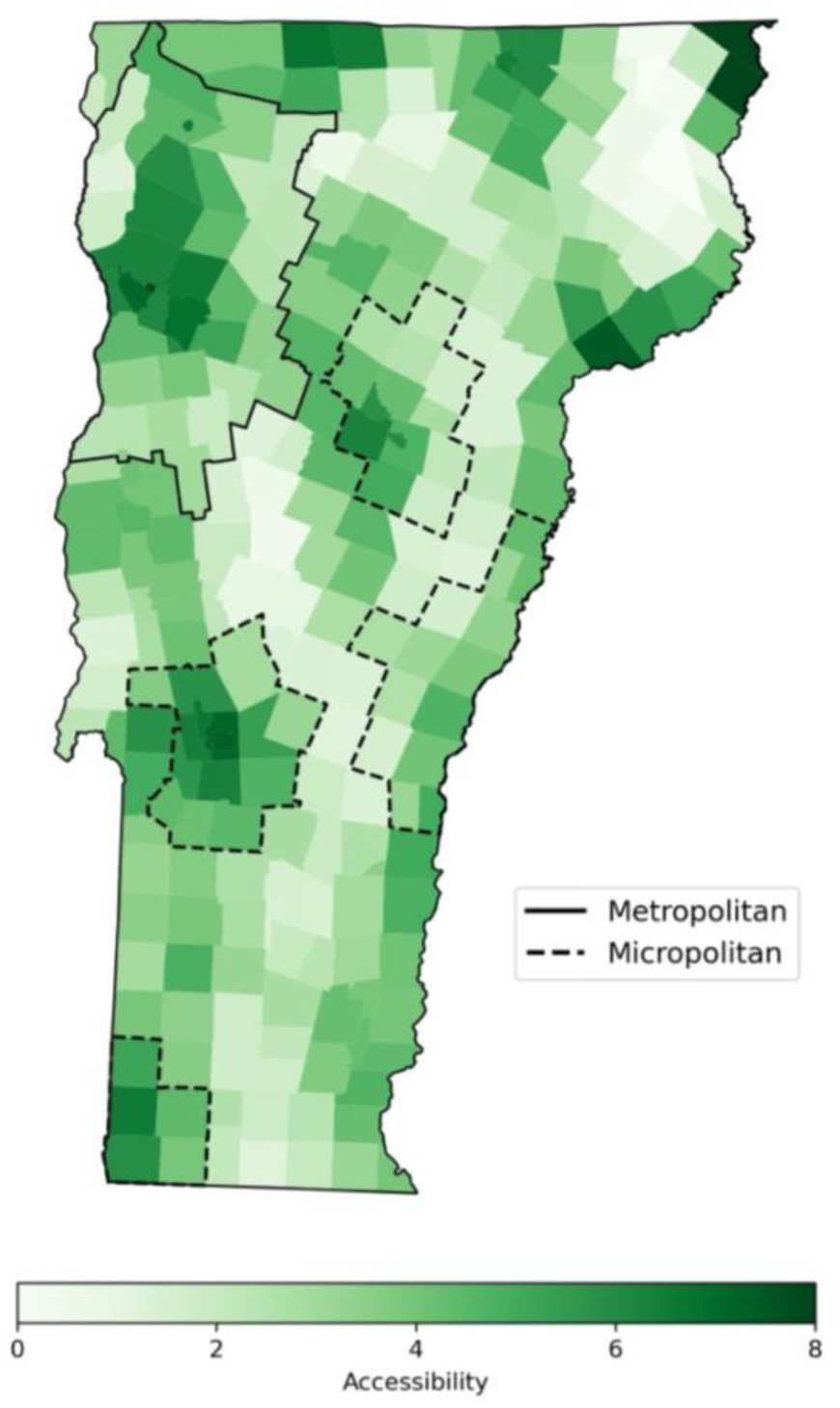
Spatial accessibility during conventional weekday business hours.

The number of available pharmacists per 10,000 residents was almost a third lower for rural areas than for metropolitan and micropolitan areas at times of peak access. Two sparsely populated rural areas in the Northeast portion of the state also demonstrated very high accessibility to pharmacies (Figure 2) due to a pair of independent pharmacies nearby in New Hampshire.

### Temporal Variation in Spatial Accessibility

Accessibility of pharmacies varies temporally over different days of the week and times of day on typical weekdays, Saturdays, and Sundays. Accessibility drops considerably weekends and outside of normal business hours. Mean peak accessibility was 3.41 on weekdays but dropped to 2.02 on Saturdays and 1.43 on Sundays. This represents a drop of 41% on Saturdays and 58% on Sundays compared to weekdays. We found that there was a significant difference in mean access between these days (Kruskal-Wallis *H* = 202.7, *p*<0.001). As accessibility drops across all areas of the state on the weekends, the overall geographic pattern of accessibility remains similar, with metropolitan and micropolitan areas sustaining more access than rural areas at all times (Figure 3).

**Figure 3.**
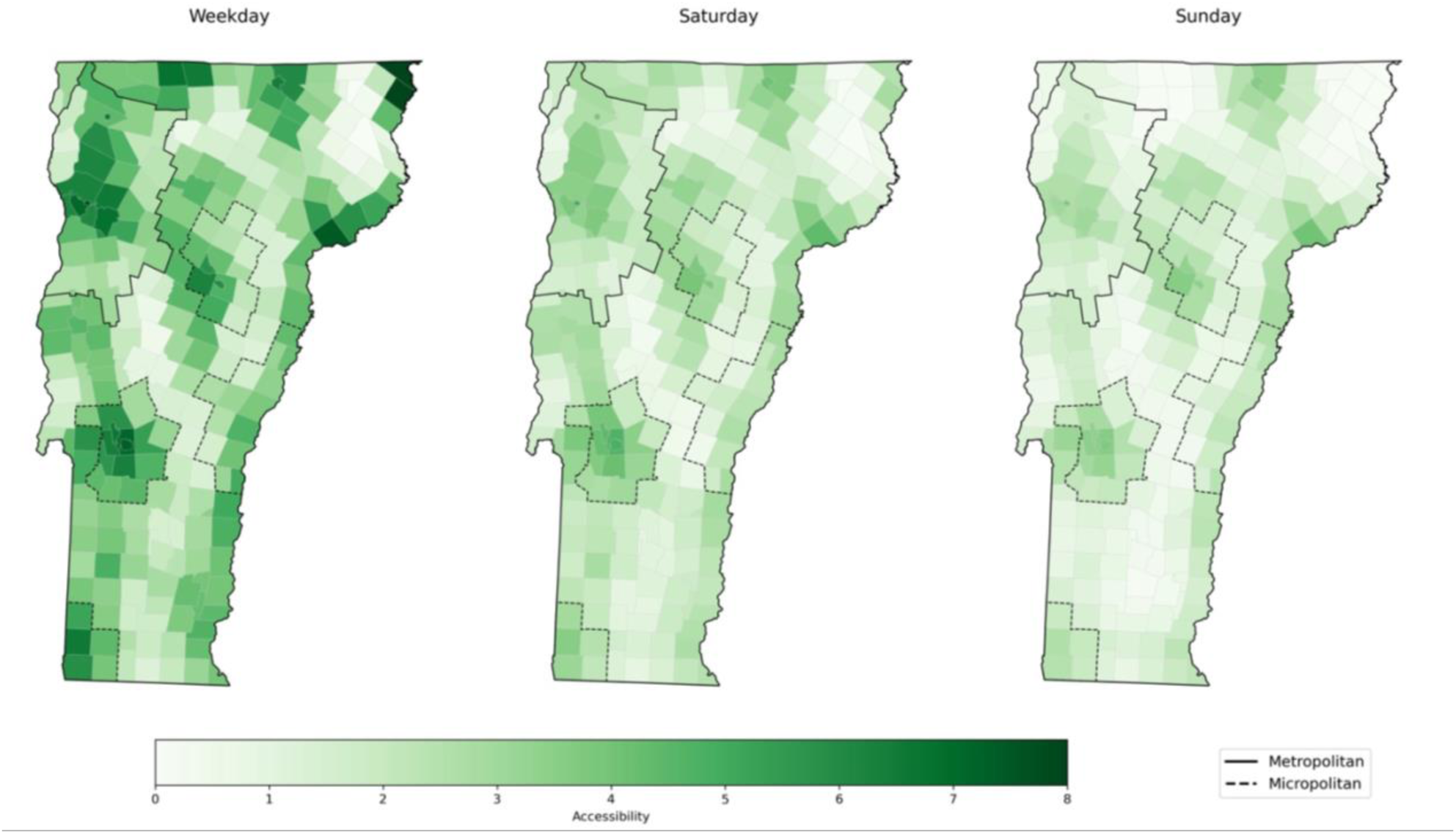
Spatial accessibility across days of the week. Each map represents the maximum accessibility on each day when all pharmacies that plan to open that day are operational (i.e., at 12 pm).

Pharmacies are more accessible earlier in the morning and later in the evening on weekdays than on weekends (Figure 4). There is almost no retail pharmacy service in Vermont before 8 am weekday mornings and 9 am weekend mornings. Access was relatively constant between 10 am and 4 pm on all days. In the evening, pharmacy services steadily close from 5 pm to 10 pm on weekdays, and close all services by 8 pm on weekends. We have not analyzed late evening and nighttime accessibility because there were no 24-hour pharmacies in Vermont. Just two out-of-state pharmacies provide 24-hour access near the southeast border of Vermont (Figure 5). For the vast majority of the state, there are no retail pharmacy services between the hours 10 pm and 7 am on any day.

**Figure 4.**
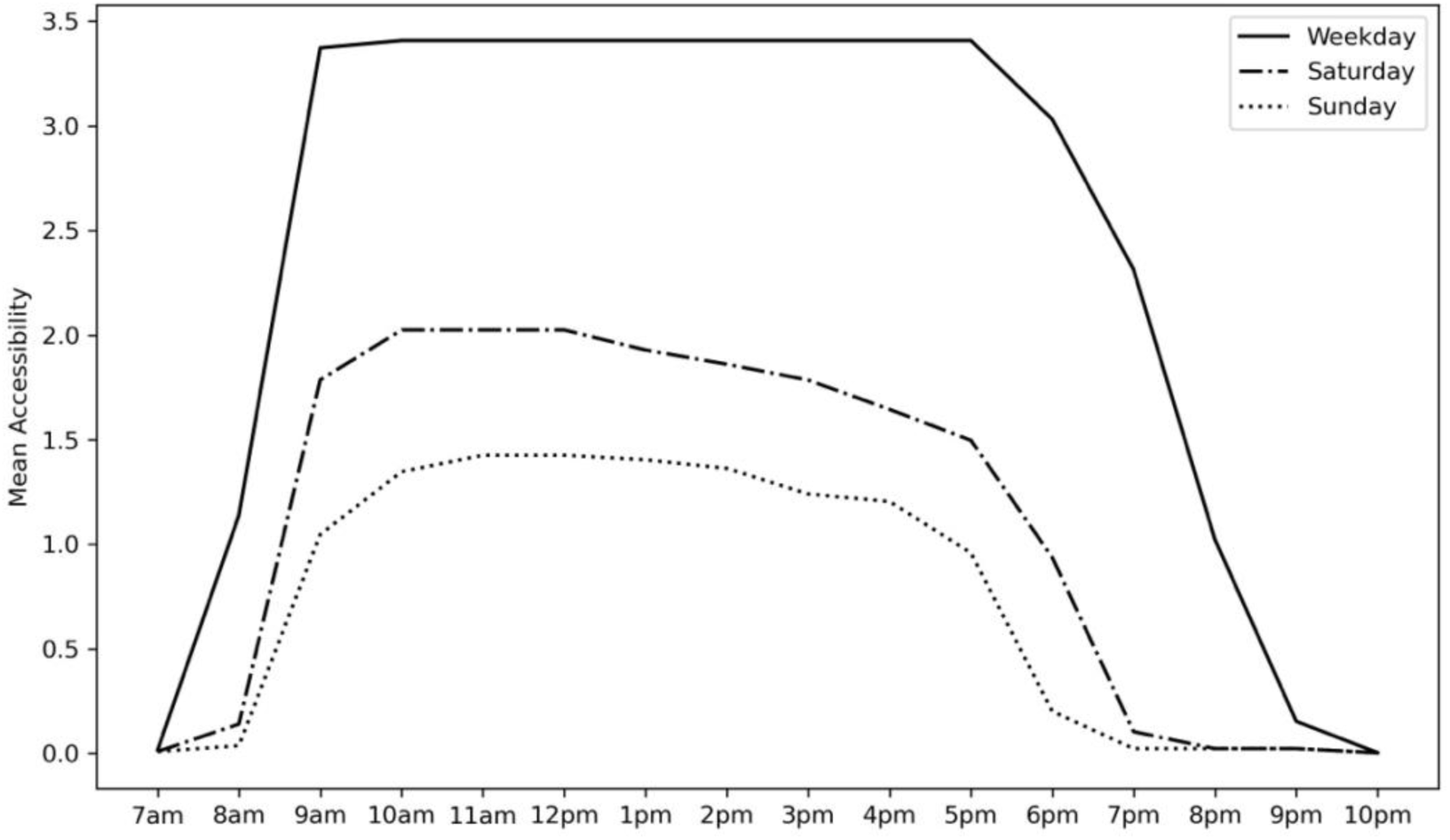
Mean spatial accessibility throughout hours of the day by days of the week.

**Figure 5.**
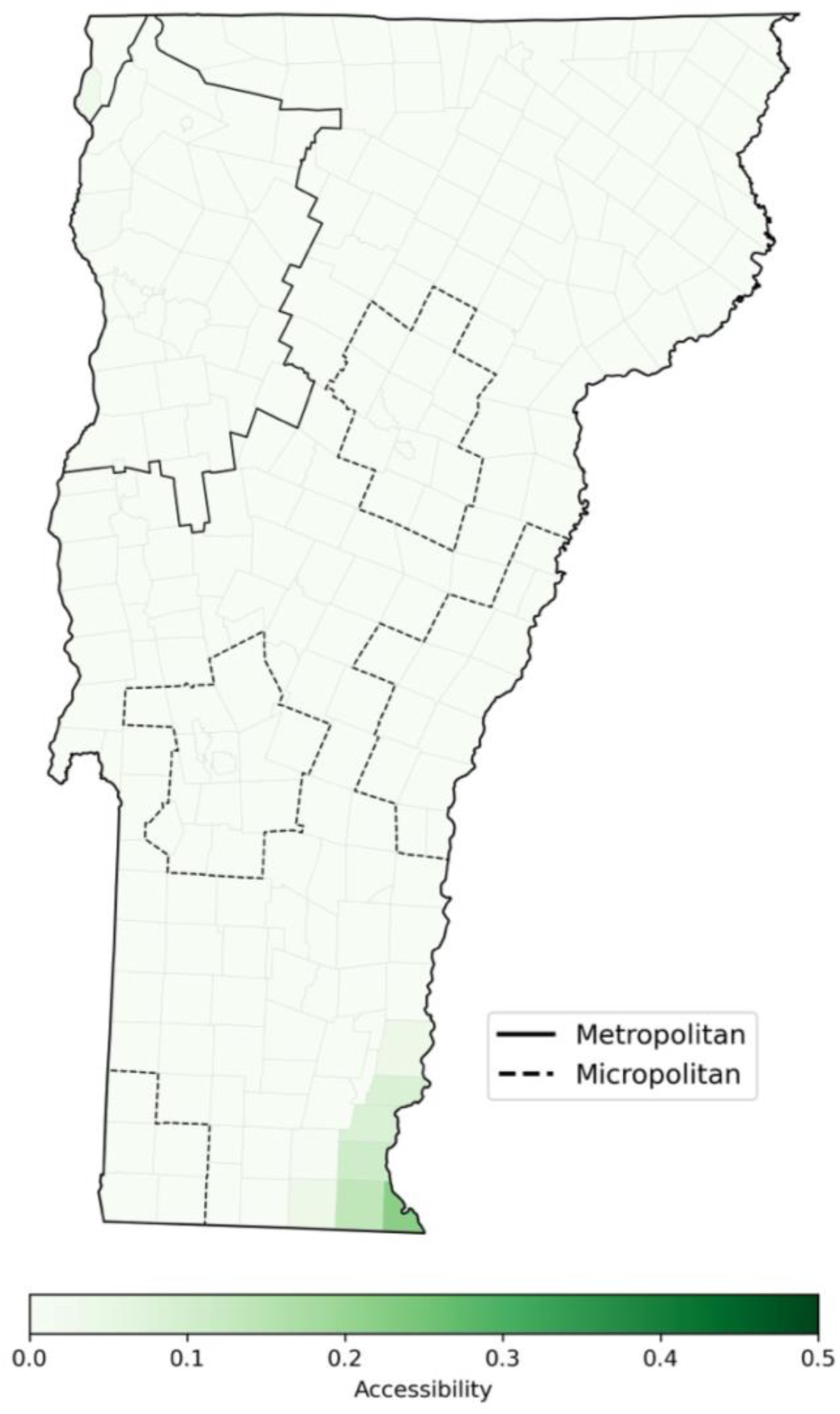
Late-night accessibility. Represents access between 10 pm and 7 am.

### Varying Spatial Differences across Temporal Segments

Finally, we found that urban-rural differences in access are greater outside of conventional business hours. Average accessibility of pharmacy care in rural areas exhibited a more pronounced decline during weekends when contrasted with urban micropolitan or metropolitan areas (Table 1). On weekdays, the average accessibility across rural towns was 28% lower than that of non-rural towns. However, accessibility across rural areas declined more severely on the weekend, dropping by 31% on Saturdays and 40% on Sundays compared to non-rural municipalities. On an hourly basis, there is a greater percent difference between rural and urban areas on the weekend than on weekdays.

**Table 1.** Non-rural versus rural mean access summary across days of the week with percent differences.

|  | *n* | *Weekday Mean*<br>*Access* | *Saturday Mean*<br>*Access* | *Sunday Mean*<br>*Access* |
| --- | --- | --- | --- | --- |
| *Non-Rural* | 83 | 4.23 | 2.56 | 1.95 |
| *Rural* | 172 | 3.01 | 1.76 | 1.18 |
| *Difference* | — | -28% | -31% | -40% |

In addition to exacerbated urban-rural differences on weekends, accessibility differences are also greater outside of conventional business hours (Figure 6), as urban pharmacies tend to open earlier and close later than rural pharmacies. The percent difference in mean accessibility between urban and rural areas is highest before 9 am and from 5 pm to 9 pm, while remaining steady during normal working hours. The difference is not meaningful from 10 pm to 6 am, when all Vermont-based retail pharmacies are closed. In sum, the differences in pharmacy accessibility between urban and rural areas are greater on weekends and at the morning and evening fringes of normal working hours.

**Figure 6.**
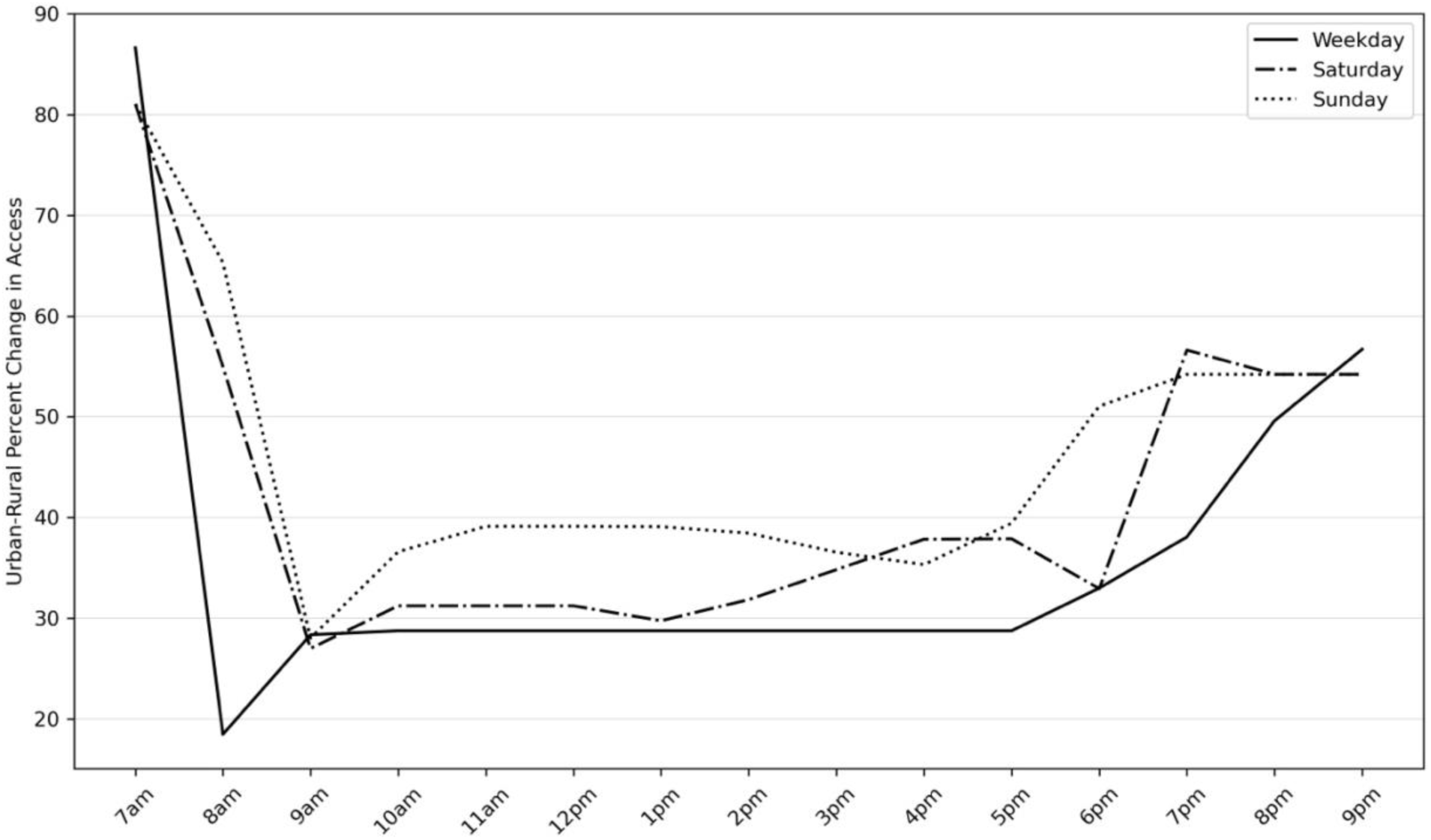
Percent difference in mean accessibility scores across hours of the day between rural and non-rural (metropolitan and micropolitan) areas by day of the week.

## Discussion

In this study, we evaluated spatiotemporal variation in access to pharmacy care across Vermont, focusing on the accessibility differences between rural, micropolitan, and metropolitan areas for different days of the week and times of day. Our results are broadly consistent with other national studies of pharmacy access. Qato et al.’s [44] county-level analysis showed that Vermont’s counties had an average number of pharmacies per population and lacked advanced services like 24-hour or multilingual services. Another study [45] of pharmacy access across the US found that of Vermont’s 184 census tracts, 37 tracts were characterized as low access and 4 tracts were characterized as pharmacy deserts. Our study improves upon this prior work and reveals new findings by incorporating 1) spatial interaction and distance decay with the E2SFCA method while accounting for boundary and edge effects, 2) varying levels of service at different pharmacy locations, 3) operating hours and their influence on spatial accessibility over time, and 4) open science research practices. These methodological improvements and the findings that they reveal are crucial in the context of rural health care and current economic restructuring in the retail pharmacy industry.

Unlike many studies of geographic access to pharmacies studies, our methods account for threats to validity rooted in geographic edge and boundary effects. At the edge of a study region like Vermont, residents may use services outside of the state and non-residents may use services inside the state. Our model therefore includes roads, pharmacy locations, and population in nearby regions of New Hampshire, Massachusetts, and New York. Most accessibility measures also simplify residential areas into points, causing analysis to be very sensitive to the configuration of residential area boundaries. We have improved upon these methods by preserving residential area polygons and calculating area-weighted estimates of the populations within each distance threshold of a service location.

We are not aware of any other 2SFCA or E2SFCA pharmacy studies that have incorporated service capacities varying by location into their models. However, accounting for variation in service levels is essential for accurately modeling real-world differences in pharmacy capacity and identifying meaningful disparities in access. For example, an independent pharmacy with specialized services and extra pharmacists and staff does not offer the same capacity as a retail chain pharmacy maximizing profits with streamlined services and minimized staffing. Service level variation may be especially important in rural areas such as Vermont, where a single under-staffed provider may represent a “keystone” pharmacy as the only point of access [10].

Our findings demonstrate that the most significant disparities in access between urban and rural towns occur on weekends and after hours, but the literature overlooks these differences by failing to account for time. Consistent with the existing literature on urban-rural disparities of pharmacy access [20,22,23], our results show consistently higher access in metropolitan areas like Burlington and lower access in rural areas. However, we also observed a dramatic drop in services on weekends and after hours across all town types. In fact, the magnitude of temporal differences exceeded that of urban-rural disparities, and the temporal differences were more extreme in rural areas than urban areas. Taken together, these findings suggest that spatial accessibility studies without a temporal dimension may miss crucial weekend and after-hours differences between urban and rural areas. Crucially, studies without a temporal dimension cannot capture a common experience in Vermont: where the only place to fulfill prescriptions after hours is a hospital emergency room.

Temporal considerations are crucial given the time-sensitive essential medications and medical services community pharmacies provide, such as emergency overdose treatment [46], emergency contraception [47], post-exposure prophylaxis [48], or filling prescriptions resulting from off-hours telehealth visits. A prior study found that 10.7% of chain pharmacies and 4.9% of all pharmacies nationally offered 24-hour services in 2015 [44]. By contrast, none of Vermont pharmacies provide 24-hour services, indicating greater difficulty accessing late-night pharmacy services than the national average. Pharmacies open 24 hours a day are critical in providing round-the-clock access to the aforementioned essential services and medications.

Our study is particularly timely and valuable in the context of the ongoing widespread closures of community pharmacies across the United States, with over 7,000 pharmacies having closed in recent years and more expected [8,9,11]. Recent research suggests that these closures could disproportionately burden rural residents and those living in medically underserved areas [10,49].

By following open science practices in this study, we have facilitated the possibility of reanalysis, replication, and broader policy impacts. With only limited python coding experience, anyone can adapt our code to adjust the measurement of pharmacy services, modify distance parameters, or substitute the population data to be used. In the context of widespread closures, the study can be replicated over time to track changes or applied in different geographic regions. Additionally, study data and parameters can be adjusted to simulate future scenarios –– such as reductions in operating hours, staffing shortages, and closures community pharmacies –– to inform policy. These impacts are made possible with open-source software, cyber-GIS infrastructure, and open code and research protocols with licenses permitting reuse.

### Limitations

We aimed to provide a fully reproducible study with complete staffing information for all locations. However, staffing levels are not publicly available, and some retail location managers were unwilling to provide this information. In conversation with managers, we also realized that there can be inconsistency and uncertainty in staffing levels, especially in the context of hiring and staffing shortages. It was therefore infeasible to model hourly staffing variation at individual pharmacies. In terms of model parameters, further research is required to refine the estimation of service levels per pharmacy, distance thresholds, and distance weights. It may be possible to use prescription fulfillment or claims data to calibrate and validate subjective modeling patterns like the rate of distance decay and the calculation of the level of services per pharmacy. As a relatively understudied dimension of healthcare access, prior studies and data sources are not readily available for determining and calibrating these study parameters. Future work is also needed to link spatial accessibility to health outcomes. However, to our knowledge, no studies of spatial accessibility to pharmacies have directly assessed these associations. This reflects a broader challenge in the field of meaningfully linking pharmacy access to specific downstream health outcomes due to the complex interplay of factors influencing health and the lack of individual-level, longitudinal data connecting pharmacy use to clinical outcomes.

We acknowledge that some studies recommend varying catchment distance parameters across urban-rural gradients in a study area to account for different distances that urban and rural populations are willing to travel for care [18,20]. Although this modification might yield better estimates of demand and service-to-population ratios in the first step of E2SFCA, it also diminishes the disproportionate travel cost borne by rural residents. We also acknowledge that measures of spatial inaccessibility may be combined with socio-demographic data to define pharmacy “deserts” [50], but this was beyond the scope of our research questions. Future research in Vermont could include socioeconomic variables to further inform intervention.

### Conclusion

We utilized the E2SFCA method to reveal important urban-rural differences in the spatial accessibility of pharmacies across Vermont and characterized how these accessibility differences vary based on temporal characteristics throughout a given week. Rural towns have considerably worse access to pharmacy care in the state compared to metropolitan and micropolitan towns, but this difference in accessibility fluctuates throughout days of the week and times of the day, further confirming that considering the temporal dimension in spatial accessibility of healthcare studies can have important public health implications. In our case, publicly accessible datasets did not suffice for investigating the spatial accessibility of pharmacies in Vermont. Instead, we implemented a data collection process that was crucial to understanding the variation in accessibility. Future spatial accessibility analyses should consider undertaking similar data collection to investigate spatial accessibility of healthcare when all necessary datasets may not be preexisting. Public centralized datasets need to include more information on operational hours and staffing or services in order to support this type of research.

Our findings highlight the need for targeted policy intervention to alleviate disparities in access to this increasingly critical part of primary healthcare. There are direct implications for service providers and healthcare planning in Vermont. By identifying disparities in pharmacy accessibility between town types and temporal segments, our analysis provides potential areas for targeted intervention to improve healthcare access for Vermont residents. Extending hours of operations and increasing pharmacy staffing on weekends across select Vermont pharmacies could substantially improve Vermont residents’ access to essential medications and healthcare services. Considering the exacerbated urban-rural accessibility disparity on weekends, it may be particularly valuable to focus on implementing these measures in Vermont’s rural regions, effectively improving access for a significant portion of Vermont towns. Policy intervention should also aim to address the complete void in accessibility during extreme hours, such as late nights or early mornings, which indiscriminately impacts all town types. Ensuring and expanding access to pharmacies is a key component of increasing accessibility to primary healthcare for Vermont municipalities and may be one component to smoothing urban-rural disparity in health outcomes.

## Declarations

### Ethics approval and consent to participate

Our research protocol was approved by the Middlebury College Institutional Review Board Protocol #264.

### Consent for publication

Not applicable

### Availability of data and materials

Our data and code are available in an OSF project: Roubin, S., Holler, J., & Kedron, P. (2024, September 18). Spatio-Temporal Accessibility of Pharmacy Care in Vermont, USA. https://doi.org/10.17605/OSF.IO/BCQ9S

The OSF project is linked to a GitHub repository https://github.com/HEGSRR/OR-VT-Pharmacy. Data on staffing levels is confidential, but a synthetic dataset with similar statistical properties is available in the repository.

### Authors and Affiliations

Department of Geography, Middlebury College, Middlebury, VT, USA Samuel Roubin & Joseph Holler

Department of Geography University of California, Santa Barbara, Santa Barbara, CA, USA Peter Kedron

### Authors’ contributions

JH, SR: conceptualization. JH: supervision. SR: data collection. SR, JH: data curation; data analysis. SR: writing – original draft. SR, JH, PK: writing – review and editing. All authors have read and approved the final manuscript.

## Data Availability

Our data and code are available in an OSF project: Roubin, S., Holler, J., & Kedron, P. (2024, September 18). Spatio-Temporal Accessibility of Pharmacy Care in Vermont, USA. https://doi.org/10.17605/OSF.IO/BCQ9S. The OSF project is linked to a GitHub repository https://github.com/HEGSRR/OR-VT-Pharmacy. Data on staffing levels is confidential, but a synthetic dataset with similar statistical properties is available in the repository.

## Acknowledgments

Not applicable.

## Abbreviations

2SFCA: Two-step Floating Catchment Area Method
E2SFCA: Enhanced Two-step Floating Catchment Area Method GIS: Geographic Information Systems
VTDOH: Vermont Department of Health

